# The Dynamic Changes of Antibodies against SARS-CoV-2 during the Infection and Recovery of COVID-19

**DOI:** 10.1101/2020.05.18.20105155

**Authors:** Kening Li, Min Wu, Bin Huang, Aifang Zhong, Lu Li, Yun Cai, Lingxiang Wu, Mengyan Zhu, Jie Li, Ziyu Wang, Wei Wu, Wanlin Li, Bakwatanisa Bosco, Zhenhua Gan, Zhihua Wang, Qinghua Qiao, Jian Wu, Qianghu Wang, Shukui Wang, Xinyi Xia

## Abstract

Deciphering the dynamic changes of antibodies against SARS-CoV-2 is essential for understanding the immune response in COVID-19 patients. By comprehensively analyzing the laboratory findings of 1,850 patients, we describe the dynamic changes of the total antibody, spike protein (S)-, receptor-binding domain (RBD)-, and nucleoprotein (N)- specific IgM and IgG levels during SARS-CoV-2 infection and recovery. Our results indicate that the S-, RBD-, and N- specific IgG generation of severe/critical COVID-19 patients is one week later than mild/moderate cases, while the levels of these antibodies are 1.5-fold higher in severe/critical patients during hospitalization (*P*<0.01). The decrease of these IgG levels indicates the poor outcome of severe/critical patients. The RBD- and S-specific IgG levels are 2-fold higher in virus-free patients (*P*<0.05). Notably, we found that the patients who got re-infected had a low level of protective antibody on discharge. Therefore, our evidence proves that the dynamic changes of antibodies could provide an important reference for diagnosis, monitoring, and treatment, and shed new light on the precise management of COVID-19.

## Introduction

Caused by the infection of severe acute respiratory syndrome coronavirus 2 (SARS-CoV-2), coronavirus disease 2019 (COVID-19) is spreading in more than 210 countries and territories around the World^1–3^. As of April 24, 2020, a total of 2,718,155 confirmed cases were reported, of which 190,636 patients died. Although its mortality is lower than SARS and MERS, the high infection rate leads to its rapid spread^4^. Approximately 100,000 confirmed cases increased every single day, extremely challenging the public health and medical service around the globe. Therefore, rapid, accurate, and timely diagnosis and monitoring of COVID-19 are urgently needed.

Currently, the diagnosis of COVID-19 is mainly based on the virus RNA load test using aquantitative real-time polymerase chain reaction (RT-PCR)^5^. However, the nucleic acid testing results are subject to many factors, including the specimen location, type, quality, and patients' condition, as well as sample storage. Thus, some COVID-19 patients will be unrecognized if only based on the virus RNA load test^6^. In consideration of the high false-negative rate of virus load detection, on March 3, 2020, the latest release of "Diagnosis and Treatment Protocol for Novel Coronavirus Pneumonia of China" added that SARS-CoV-2 specific IgM and IgG antibody levels could be used to diagnose the suspected cases. The antibody detection is more simple and faster than the virus RNA load test, and the specimen is more stable and easier to store^7^. Thus, antibody tests can provide an important complementary for the diagnosis of COVID-19.

In addition, the generation and maintenance of neutralizing antibodies against SARS-CoV-2 play an important role in resisting the virus infection by host^8^. It is widely recognized that IgM provides the first line of defense during viral infections, while the production of IgG lags behind IgM, and provides long-term immunity and memory. The coronavirus neutralizing antibodies primarily target the trimeric spike (S) glycoproteins on the viral surface that mediates entry into host cells^9^. The S protein has two functional subunits, the S1 subunit mediates cell attachment, and the S2 subunit mediates fusion of the viral and cellular membrane^10^. Neutralizing antibodies often target the receptor-binding domain (RBD) in the S1 subunit to block the interaction between the virus and host receptor^11^. However, the dynamics of serum antibodies against SARS-CoV-2 during COVID-19 infection and recovery are still unclear, and most of the current opinions are based on the knowledge acquired from previous experience of the SARS-CoV and MERS-CoV infections^12^.

Here, we comprehensively analyzed the laboratory tests of 1,850 hospitalized COVID-19 patients at Wuhan Huoshenshan Hospital, admitted from February 4 to March 30. We describe the dynamics of the SARS-Cov-2-specific antibody levels, including total antibody, as well as the S-, RBD-, and nucleoprotein (N)-specific IgM and IgG levels on admission, during hospitalization and on discharge. Our results would provide an important reference for the understanding of immune response after SARS-Cov-2 infection, and the function of protective antibody during the recovery, and shed new light on the precise management of COVID-19.

## Methods

### Data Collection

We analyzed the laboratory test results of 1,850 COVID-19 patients admitted from February 4 to March 30 at Wuhan Huoshenshan Hospital, which was one of the biggest designated hospitals for COVID-19 in Wuhan. This cohort included 796 mild or moderate cases and 1054 severe or critical cases. Information on the clinical characteristics and laboratory findings of all patients was extracted from the hospital electronic medical records. This study was approved by the Medical Ethical Committee of Wuhan Huoshenshan Hospital. Written informed consent was obtained from each patient.

### Serum Anti-SARS-CoV-2 Antibodies Assay

Total SARS-CoV-2 IgM or IgG in the serum was measured by chemiluminescence using commercially available kits (Shenzhen YHLO Biotech Co., Ltd.) in 1,850 patients at different time points. The magnetic beads of this kit are coated with recombinant N and S proteins. 416 of these patients were tested for S-, RBD-, and N-specific IgM and IgG levels at different time points by chemiluminescence using commercially available kits (Nanjing RealMind Biotech Co., Ltd.), including 126 mild or moderate patients, and 290 severe or critical patients. Briefly, blood samples were centrifugated at room temperature and the supernatant was removed and incubated separately with SARS-CoV-2 antigens-coated magnetic beads. The antigen-antibody complex captured by the beads slurry was gently precipitated by a magnetic separation rack. The beads were then incubated with acridinium ester-labeled mouse anti-human IgM or IgG antibody and reacted with hydrogen peroxide in excitation buffer. Relative luminescence intensity was recorded in an ACL2800 chemiluminescence system (Nanjing RealMind Biotech Co., Ltd.). Relative luminescence intensity was converted to antibody level and its unit is AU/mL.

### Statistical Analysis

Continuous and categorical variables were presented asmedian (IQR) and n (%), respectively. We used the two-sided Wilcoxon rank-sum test or Fisher’s exact test to compare the difference between groups where appropriate.

## Results

### 1. Temporal profiles of antibodies against SARS-CoV-2 of mild/moderate and severe/critical COVID-19 patients

To explore the temporal dynamics of immune response after SARS-Cov-2 infection, we analyzed the antibody levels at different time points after symptoms onset, and the timing and level were compared between mild/moderate and severe/critical COVID-19 patients.

The level of total IgM was extremely low in the first week (median=5.34 AU/ML), and gradually increased until the 5th week (median=43.98 AU/ML), followed by a continuous decrease to the initial level. The level of total IgG was more detectable than IgM at the first week, and continuously increased to seventh week (median=154.54 AU/ML), and slightly decreased from the 8th week, but still kept considerably high level until the end of our observation (12th week, median=95.94 AU/ML) (Supplementary Figure S1A). We further quantified the total antibody levels of confirmed patients after disease onset, and found that the positive rates of the total IgG were 62.5% in the first week. Total IgG could be detected in 94.7% of patients after 5 weeks of disease onset, and 97.6% of patients retained considerable abundance even after 12 weeks. While IgM was rarely detected (32.5%) during the early stage (SupplementaryFigure S1B). 97.4% of the confirmed patients have positive IgM or IgG at the first 1 week after symptom onset, indicating that the combination of IgM and IgG is necessary for auxiliary diagnosis.

As shown in Figure S1C, the level of S-, RBD-, and N-specific IgM reached a high level in the second week after symptom onset in the mild/moderate COVID-19 patients, while it took 3 weeks for the severe/critical COVID-19 patients to get the comparable antibody level. Furthermore, in the mild/moderate COVID-19 patients, the level of S-, RBD-, and N-specific IgG generated at the first 2 weeks after symptom onset, then sharply increased in the third week, and reached their peaks at the fifth or sixth week. However, in the severe/critical COVID-19 patients, the level of S-, RBD-, and N-specific IgG generated in the third week after symptom onset and reached their peaks in the seventh week. These IgG levels kept considerably high level until the end of our observation (12thweek). These results indicated the immune response of the severe/critical COVID-19 patients was later than the mild/moderate ones for approximately one week. We found that the S-, RBD-, and N-specific IgG levels were higher in the mild/moderate COVID-19 patients at the early stage of infection (first 2~3 weeks after onset) than the severe/critical COVID-19 patients, suggesting the lack of sufficient protective antibodies in patients who had more severe symptoms at the disease onset. But at the middle and late stages of infection (7~10 weeks after onset), the IgG levels were significantly higher in the severe/critical COVID-19 patients than the mild/moderate ones (*P*<0.05). We compared the total antibody, as well as the S-, RBD-, and N- specific antibody levels between the cured mild/moderate and severe/critical COVID-19 patients at the time of admission, hospitalization, and discharge. Results showed that total IgG, S-, RBD-, and N-specific IgG levels of the severe/critical COVID-19 patients were lower than that of the mild/moderate patients on admission, but these levels sharply increased during hospitalization and on discharge (Figure 1, Table 1). The RBD-specific IgG level was approximately 1.5-fold higher in the severe/critical COVID-19 patients than in the mild/moderate ones (*P*=0.006) during hospitalization, and this ratio is up to 1.8 on discharge (*P*=0.001). The S-specific IgG levels were also significantly higher in the severe/critical COVID-19 patients during hospitalization and on discharge (*P*<0.01, Table 1). The significant increase levels of protective antibody against SARS-Cov-2 implied that there are more activated immune responses during the recovery of the severe/critical COVID-19 patients.

**Figure 1.**
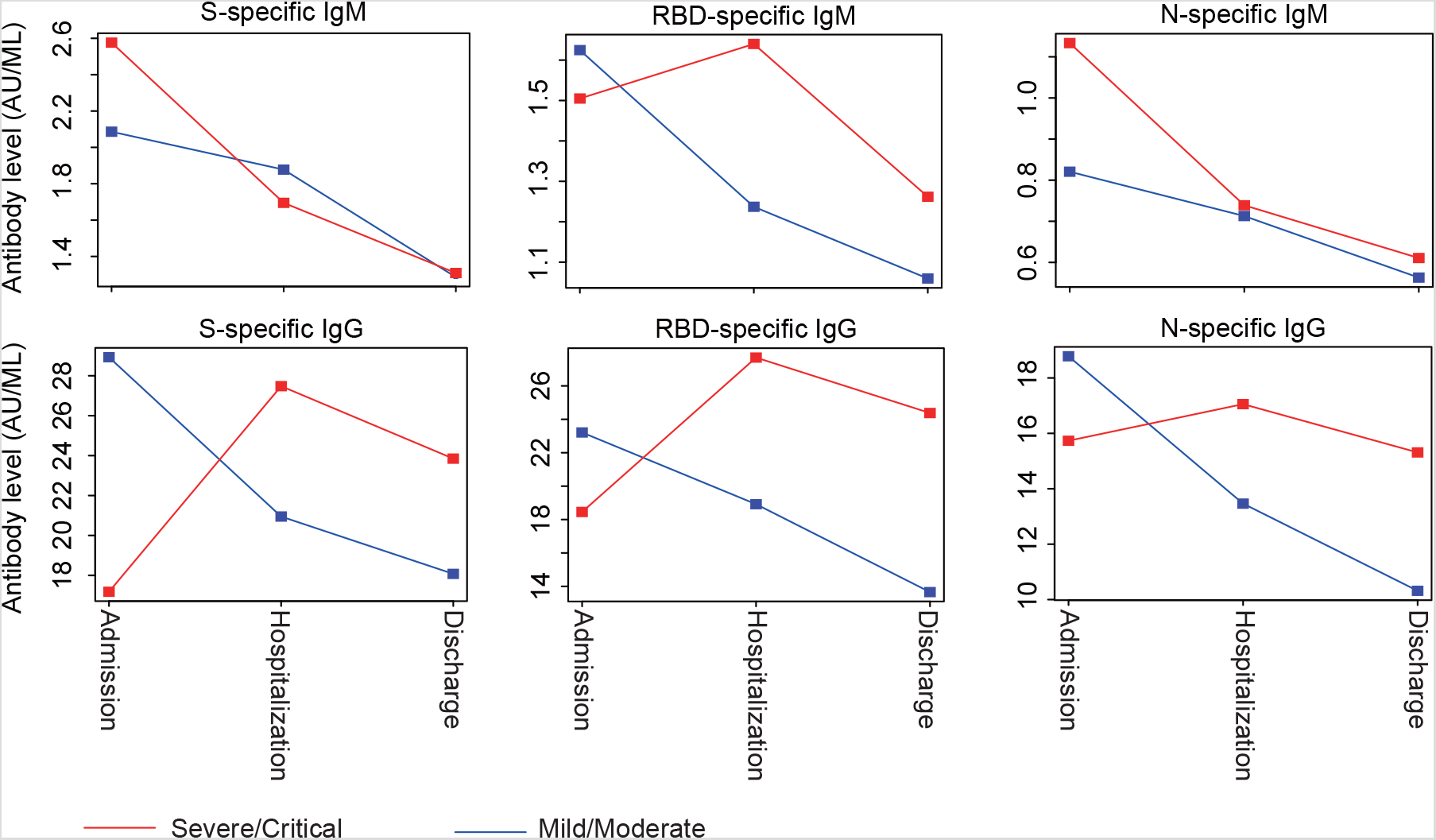
**The temporal dynamics of the S-specific, RBD-specific and N-specific IgM and IgG levels in the mild/moderate and severe/critical COVID-19 patients**. The median of S-specific, RBD-specific and N-specific IgM and IgG levels of the mild/moderate and severe/critical patients on admission, during hospitalization and on discharge.

Taken together, this evidence indicated that compared with the mild/moderate ones, the severe/critical COVID-19 patients had a late response at the beginning but stronger defense at the middle and late stage of infection. These results emphasized the crucial role of timely medical intervention for the severe/critical COVID-19 patients.

### 2. The IgG levels are significantly higher in older COVID-19 patients during hospitalization

We further evaluated the differences in antibody levels among patients of different age groups. As shown in Table 2, the patients were divided into three groups based on their age, young (Age<40 years), middle-aged (40-65 years), and older patients (Age>65 years). On admission, though not statistically significant, the RBD-specific antibody level in patients who were older than 65 years was relatively lower than in young and middle-aged patients, while the N-specific antibody levels of older patients were higher, probably because N protein had high immunogenic activity and was abundantly expressed during the early stage of infection, while the level of protective RBD-specific antibody was lower in older patients owing to their weak immune defense system. Besides, we observed that the S-, RBD-, and N-specific IgG levels were gradually elevated along with the age increase during hospitalization and on discharge (*P*<0.05). For example, the median level of RBD-specific antibody in young, middle-aged and older patients was 7.7 AU/ML, 22.4 AU/ML, and 30.7 AU/ML, respectively (young vs. middleage: *P*<0.0001, middle-age vs. older: *P*=0.003) during hospitalization. Because the older COVID-19 patients were usually more severe^1^, the more activated immune defense was observed in these patients.

### 3. The dynamic changes of the IgG levels are related to the clinical outcome of the severe/critical COVID-19 patients

As the S-, RBD-, and N-specific IgG levels significantly increased in the severe/critical COVID-19 patients during hospitalization, we attempted to evaluate the function of these antibodies in the recovery of the severe/critical COVID-19 patients. We compared the dynamics of these IgG levels between cured and dead severe/critical COVID-19 patients (Figure 2A). Results showed that the S-, RBD-, and N-specific IgG levels gradually increased from admission to discharge of survivors. However, the S- and RBD-specific IgG levels sharply decreased in the fifth week after admission in non-survivors, although these IgG reached a higher level than the survivor before the fifth week. The N-specific IgG level was much higher in the non-survivors than survivors on admission, but it gradually increased in the survivors during hospitalization, while sharply decreased in the non-survivors during hospitalization. These IgG levels showed greater fluctuation during the hospitalization of non-survivors. The Gini indexes of S-, RBD-, and N-specific IgG levels were 0.27, 0.47, and 0.16, respectively, in non-survivors. While in the survivors, these values were 0.08, 0.11, and 0.08, respectively. These results suggested that the S-, RBD-, and N-specific IgG played important roles in helping the severe/critical COVID-19 patients recover, and the decrease of these antibody levels may indicate the poor outcome. The patient's disease progression can be monitored by tracking the dynamic changes in the antibody level in COVID-19 patients.

**Figure 2.**
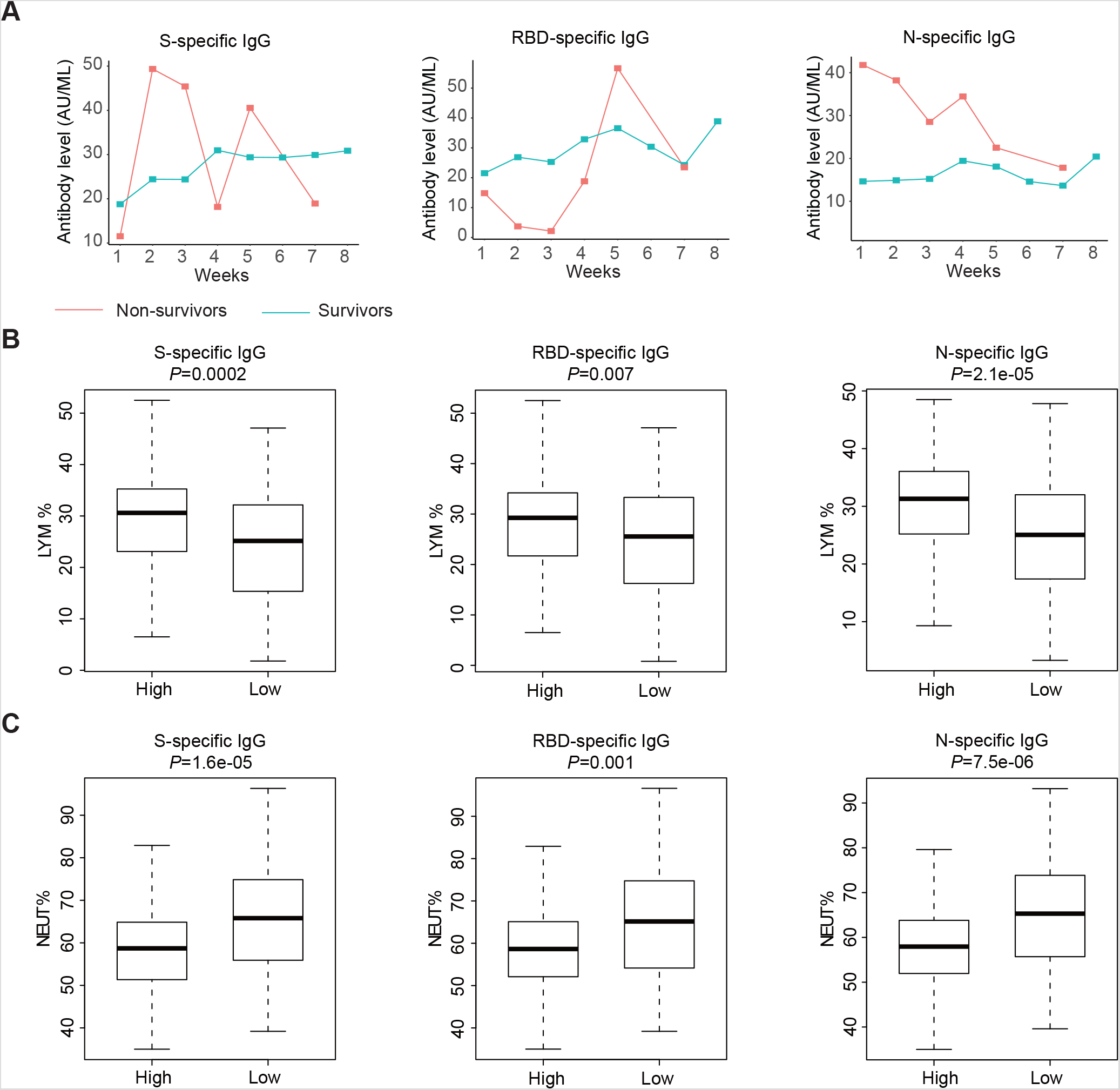
**The relationship between the outcome of the severe/critical COVID-19 patients and the dynamics of the S-specific, RBD-specific and N-specific IgG levels**. (A) The dynamics of S-specific, RBD-specific and N-specific IgG levels from admission to discharge of cured and dead severe/critical COVID-19 patients. Turquoise lines profiled the dynamics of median IgG level in survivor, and red lines profiled the dynamics of median IgG level in non-survivor. (B) The percentage of lymphocytes in patients with different N-specific, RBD-specific and S-specific IgG levels. (C) The percentage of neutrophils in patients with different N-specific, RBD-specific and S-specific IgG levels.

According to the latest research, the ratio between neutrophil and lymphocyte percentage is an important index for the prognosis of COVID-19^13^. Patients with lower lymphocyte percentage and higher neutrophil percentage often have a poor outcome^13^. To evaluate whether the antibody levels are correlated with the neutrophil and lymphocyte percentage, we analyzed the blood routine examination and antibody test data of the same cured severe/critical COVID-19 patients on the same day (Figure 2B). We observed that the percentage of lymphocytes was significantly lower in patients with a low IgG level than in patients with high IgG levels (Figure 2, S-specific IgG: *P*=0.0002, RBD-specific IgG: *P*=0.007, N-specific IgG: *P*=2.1e-05). Nevertheless, a higher percentage of neutrophils was observed in patients with low IgG levels (S-specific IgG: *P*=1.6e-05, RBD-specific IgG: *P*=0.001, N-specific IgG: *P*=7.5e-06). These results further revealed that the S-, RBD-, and N-specific IgG could prevent severe/critical COVID-19 patients from progress.

### 4. S- and RBD-specific IgG levels are significantly higher in virus-free patients

To illustrate the relationship between SARS-COV-2 virus load and antibody levels, we analyzed the test results of patients who tested both virus load and antibody level on the same day and compared the S-, RBD-, and N-specific antibody levels between virus-positive and virus-negative status. We observed that the S-specific and RBD-specific antibody levels were significantly higher in the virus-negative status than in thevirus-positive status (Table 3). The median of S-specific IgG level was 12.8 AU/ML vs. 25.1 AU/ML between virus-positive and virus-negative status (*P*=0.07), and the median of RBD-specific IgG level was 13.3 AU/ML vs. 28.3 AU/ML between virus-positive and virus-negative status (*P*=0.03), indicating the important role of protective antibodies in virus clearance.

### 5. The dynamic changes of antibody levels after convalescent plasma transfusion (CPT) therapy

Because the plasma of patients who have recovered from infections contains theneutralizing antibodies against the virus, human CPT is an option for the prevention and treatment of COVID-19 disease and has been applied in China^14,15^. We analyzed the total antibody level in patients who received CPT at different time points in our cohort. The total IgM and IgG level of 5 patients at 3 days before CPT, and 1-3, 4-7, 7-14 days after CPT were showed in Figure 3. The results showed that although some fluctuations were observed in 2 patients, the total IgM levels decreased within 14 days after CPT. While the total IgG levels decreased during the first 7 days and increased eventually in the second week. These observations demonstrate that CPT therapy may provide the long-term antibody against the SARS-Cov-2 virus for patients and help the COVID-19 patients recover. However, the molecular mechanism of the decrease of IgG level in the first week after CPT needs to be further illuminated.

**Figure 3.**
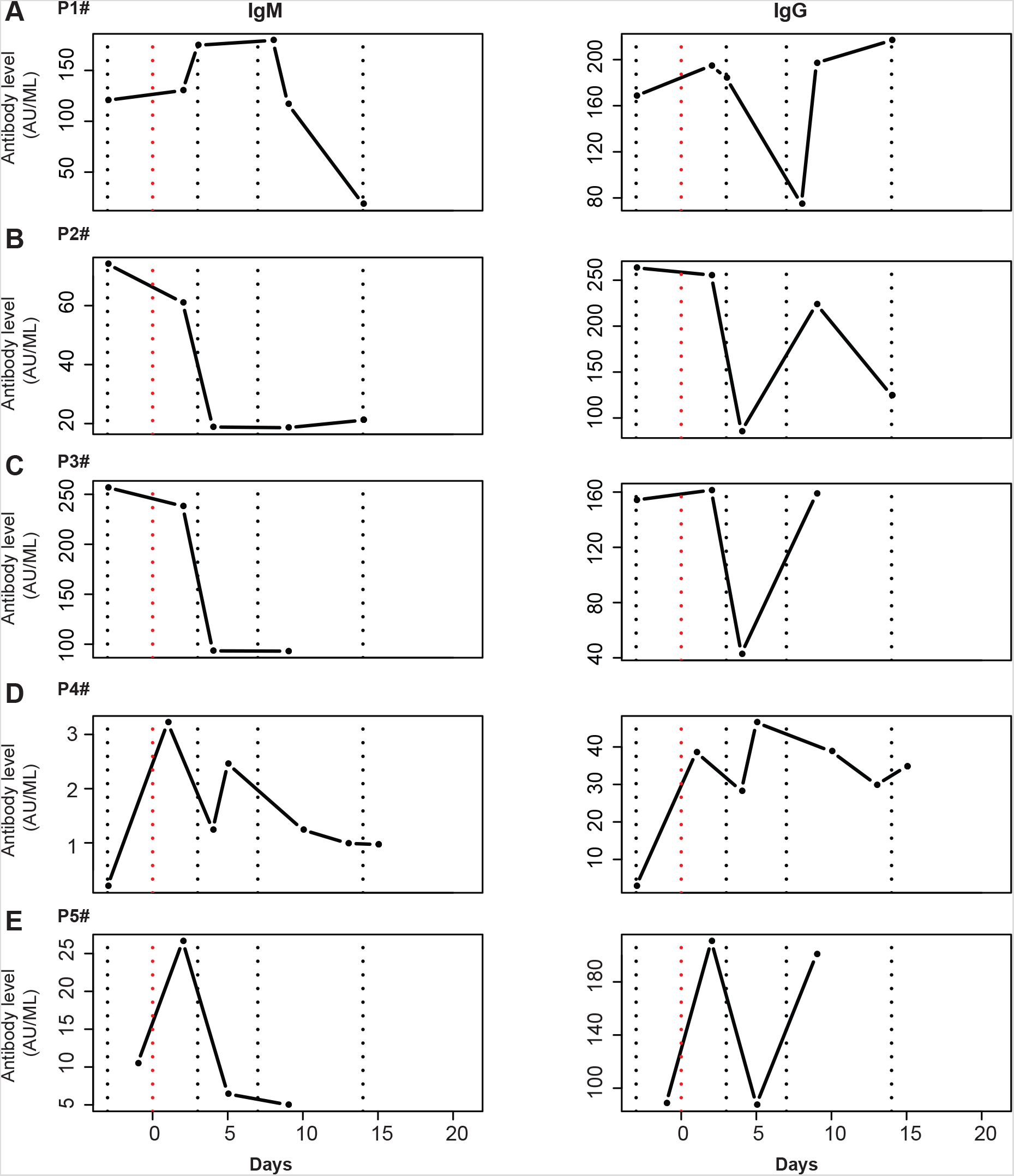
**The dynamic changes of total IgG and IgM level after convalescent plasma transfusion therapy in 5 patients**. The x-axis represents the days after convalescent plasma transfusion therapy, and y-axis represents the antibody level. Red dash line represents the day of convalescent plasma transfusion. Blue dash lines represent −3, 3, 7 and 14 days after convalescent plasma transfusion, respectively.

### 6. The low level of protective antibody on discharge is associated with a high risk of re-infection

According to the latest version of "Diagnosis and Treatment Protocol for Novel Coronavirus Pneumonia of China", patients who meet the following criteria can be discharged: 1) body temperature is back to normal for more than three days; 2) respiratory symptoms improve obviously; 3) pulmonary imaging shows obvious absorption of inflammation; 4) nucleic acid tests negative twice consecutively on respiratory tract samples such as sputum and nasopharyngeal swabs (sampling interval being at least 24 hours). However, we are wondering whether the discharged patients have an efficient protective antibody to prevent them from re-infection. We observed that some patients tested free of virus load, but harboured low levels of protective antibody (Figure 4). In our cohort, patient #288 was discharged on March 5, 2020, with a relatively low level of S- (15.1 AU/ML), RBD- (19.2 AU/ML), and N-specific (15.9 AU/ML) antibodies, which was much lower than average level. This patient got re-infected on March 15 and was back to the hospital for treatment. His second time of discharge was on April 2, still with a low level of protective antibody. Another 3 patients who also got re-infected, their antibody levels were not tested at their first-time discharge. On their second time of discharge, the test data showed that they still had a low level of protective antibody, implying the probability that these patients were at high risk of re-infection. Therefore, our results indicated that a fair number of virus-free patients were discharged with a low level of protective antibody. These patients may need close monitoring after discharge.

**Figure 4.**
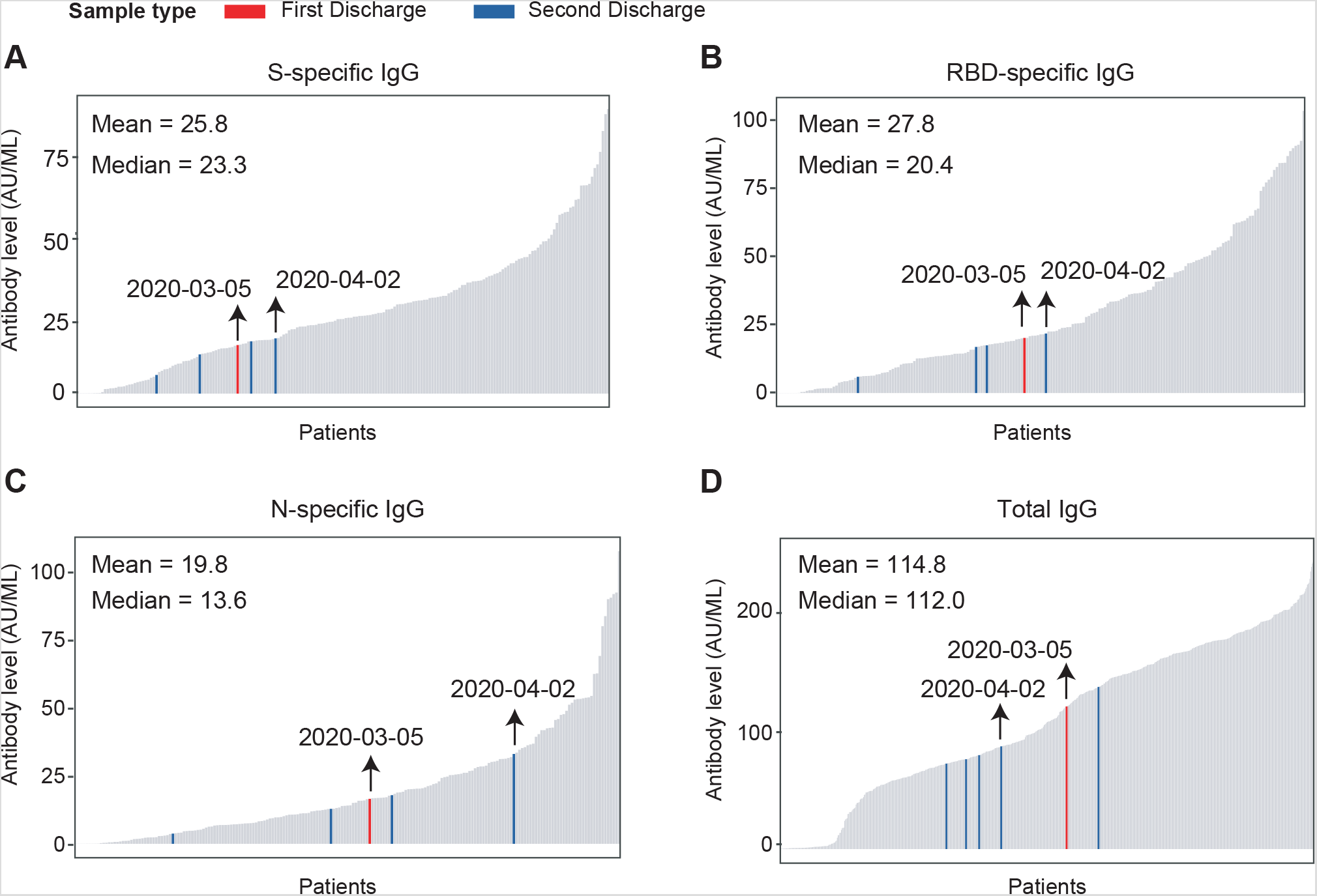
**The level of IgG on the day of discharge**. Barplot displays the level of S-specific (A), RBD-specific (B), N-specific (C) and total IgG (D) in patients on the day of discharge. The red bar indicates the first time of discharge, and blue bar indicates the second time of discharge of re-infected patients. The black arrows indicate the antibody levels of patient #288.

## Discussion

Although several case series about the SARS-Cov-2 antibody responses were previously reported^16–18^, the general pattern of the dynamics of serum anti-SARS-CoV-2 antibody during the infection and treatment of COVID-19 was still unclear due to insufficient sample size of test data. By analyzing the laboratory data of patients from Wuhan Huoshenshan Hospital, which is one of the biggest designated hospitals for COVID-19 in Wuhan, we profiled the temporal dynamic changes of the antibody level from disease onset to 12 weeks, including the S-, RBD-, and N-specific antibodies. We found that compared with the mild/moderate ones, the S-, RBD-, and N-specific IgG generated one week later in the severe/critical COVID-19 patients. These IgG levels are significantly higher in the patients who are more severe or older during hospitalization, indicating that the severe and older patients have more activated immune defense during recovery. The increase of these IgG levels indicates a better prognosis of the severe/critical COVID-19 patients. In addition, we found that the RBD-specific IgG level was much higher in virus-negative COVID-19 patients, indicating an important role of the neutralizing antibodies in virus clearance. Importantly, the patients who got discharged with low levels of protective antibody are found to be at high risk of re-infection, indicating the prognostic role of antibody level for discharged COVID-19 patients. Overall, our results suggested that these IgG, especially S-specific or RBD-specific IgG played an important role in virus clearance and recovery of COVID-19 patients.

Furthermore, according to the previous report about SARS in 2003, IgM could be detected in patients’ blood after 3-6 days of disease onset, while IgG could be detected after 8 days of disease onset^17^. Our observations showed that the total anti-SARS-Cov-2 IgG level was already at a relatively high level in the first week after disease onset, probably because some COVID-19 patients are asymptomatic at the beginning of disease onset^19,20^. The clinicians record the first day of the patients' symptoms, such as fatigue, fever, cough or diarrhea as the disease onset time point. However, the record date may be later than the actual date of infection due to the asymptomatic situation of COVID-19, so that the observed IgG level was high during the first week after record onset.

Moreover, some re-infected patients were reported lately^21,22^, attracting attention to the patients' condition on discharge. The current discharge criteria are mainly based on virus clearance and radiologic findings. However, our study shows that those re-infected patients were discharged with a low protective antibody level, emphasizing that antibody tests are necessary when discharging patients. In our opinion, two reasons may be involved in the re-infection of COVID-19 patients. Firstly, studies have shown that SARS-CoV-2 mainly infects the lower respiratory tract^23^, but the collection of the bronchoalveolar lavage requires skilled operators and specific devices, exposing the clinicians to a high risk of infection. So, the nasopharyngeal swab samples are usually used to assess the virus load, which easily leads to the false negative of discharged patients. Secondly, the immune defense system of some discharged patients is relatively weak. Lacking sufficient protective antibody makes these patients infected again. Thus, it is important to test the S-specific or RBD-specific IgG level before discharging patients and keep close monitoring of the patients with a low level of protective antibody.

In conclusion, our results suggest that the protective antibodies against SARS-CoV-2 play an essential role in COVID-19 recovery. Tracking the dynamic changes of these antibodies can provide important reference for diagnosis, monitoring, and prognosis of COVID-19, and shed new light on the development of novel agents for COVID-19.

## Data Availability

All the data supporting the findings of this study are available within the article and its Supplementary information files and from the corresponding author upon reasonable request. A reporting summary for this article is available as a Supplementary information file.

## Acknowledgements

This study was supported by the National Natural Science Foundation of China (Grant Nos. 81572893, 81972358, 81959113), Key Foundation of Wuhan Huoshenshan Hospital (Grant No. 2020[18]), Key Research & Development Program of Jiangsu Province (Grant Nos. BE2017733, BE2018713), Medical Innovation Project of Logistics Service (Grant No. 18JS005), Basic Research Program of Jiangsu Province (Grant No. BK20180036).

## Author contributions

Xinyi Xia, Shukui Wang, and Qianghu Wang had full access to all of the data in the study and takes responsibility for the integrity of the data and the accuracy of the data analysis. Kening Li, Min Wu, Bin Huang, and Aifang Zhong contributed equally. Concept and design: Xinyi Xia, Shukui Wang, and Qianghu Wang. Data collection: Aifang Zhong, Zhenhua Gan, Zhihua Wang, Qinghua Qiao, Jian Wu. Data analysis and interpretation: Kening Li, Min Wu, Bin Huang, Lu Li, Yun Cai, Lingxiang Wu, Mengyan Zhu, Jie Li, Ziyu Wang, Wei Wu, Wanlin Li. Drafting of the manuscript: Kening Li, Min Wu, and Bakwatanisa Bosco.

## Competing interests

We declare no competing interests.

**Supplementary Figure S1. The temporal dynamics of theantibody levels in COVID-19 patients after infection**. (A) The total IgM and IgG level of COVID-19 patients. The x-axis displays the weeks after symptom onset, and y-axis displays the level of total IgM and IgG. Each boxplot depicts the level of antibody, and whiskers represent the maximum and minimum. Red line based on median is used to profile the variation tendency. (B) The positive rate according to the total IgM and IgG level. The x-axis is the weeks after symptoms onset, and the y-axis shows the antibody-positive ratio of confirmed patients. (C) The dynamics of S-specific, RBD-specific and N-specific IgM and IgG levels of COVID-19 patients. The x-axis displays the weeks after symptom onset, and y-axis displays the level of antibody. Each boxplot depicts the level of antibody in 126 mild/moderate patients, and 290 severe/critical patients, and whiskers represent the maximum and minimum. Red line based on median is used to profile the variation tendency of the severe/critical patients, and blue line based on median is used to profile the variation tendency of the mild/moderate patients.

## Reference

1 Guan, W. J. et al. Clinical Characteristics of Coronavirus Disease 2019 in China. N Engl J Med, doi:10.1056/NEJMoa2002032 (2020).

2 Wu, Z. & McGoogan, J. M. Characteristics of and Important Lessons From the Coronavirus Disease 2019 (COVID-19) Outbreak in China: Summary of a Report of 72314 Cases From the Chinese Center for Disease Control and Prevention. JAMA, doi:10.1001/jama.2020.2648 (2020).

3 Grasselli, G. et al. Baseline Characteristics and Outcomes of 1591 Patients Infected With SARS-CoV-2 Admitted to ICUs of the Lombardy Region, Italy. JAMA, doi:10.1001/jama.2020.5394 (2020).

4 To, K. K. et al. Temporal profiles of viral load in posterior oropharyngeal saliva samples and serum antibody responses during infection by SARS-CoV-2: an observational cohort study. Lancet Infect Dis, doi:10.1016/S1473-3099(20)30196-1 (2020).

5 Wolfel, R. et al. Virological assessment of hospitalized patients with COVID-2019. Nature, doi:10.1038/s41586-020-2196-x (2020).

6 Li, X., Geng, M., Peng, Y., Meng, L. & Lu, S. Molecular immune pathogenesis and diagnosis of COVID-19. Journal of Pharmaceutical Analysis (2020).

7 Li, Z. et al. Development and clinical application of a rapid IgM IgG combined antibody test for SARS-CoV-2 infection diagnosis. Journal of medical virology (2020).

8 Jiang, S., Hillyer, C. & Du, L. Neutralizing Antibodies against SARS-CoV-2 and Other Human Coronaviruses. Trends in Immunology (2020).

9 Du, L. et al. MERS-CoV spike protein: a key target for antivirals. Expert opinion on therapeutic targets 21, 131–143 (2017).

10 Bosch, B. J., van der Zee, R., de Haan, C. A. & Rottier, P. J. The coronavirus spike protein is a class I virus fusion protein: structural and functional characterization of the fusion core complex. Journal of virology 77, 8801 -8811 (2003).

11 Tai, W. et al. Characterization of the receptor-binding domain (RBD) of 2019 novel coronavirus: implication for development of RBD protein as a viral attachment inhibitor and vaccine. Cellular & molecular immunology, 1–8 (2020).

12 Prompetchara, E., Ketloy, C. & Palaga, T. Immune responses in COVID-19 and potential vaccines: Lessons learned from SARS and MERS epidemic. Asian Pac J Allergy Immunol 38, 1–9, doi:10.12932/AP-200220-0772 (2020).

13 Liu, Y. et al. Neutrophil-to-lymphocyte ratio as an independent risk factor for mortality in hospitalized patients with COVID-19. Journal of Infection (2020).

14 Casadevall, A. & Pirofski, L. A. The convalescent sera option for containing COVID-19. J Clin Invest 130, 1545-1548, doi:10.1172/JCI138003 (2020).

15 Shen, C. et al. Treatment of 5 Critically Ill Patients With COVID-19 With Convalescent Plasma. JAMA, doi:10.1001/jama.2020.4783 (2020).

16 Jin, Y. et al. Diagnostic value and dynamic variance of serum antibody in coronavirus disease 2019. Int J Infect Dis 94, 49–52, doi:10.1016/j.ijid.2020.03.065 (2020).

17 di Mauro, G., Cristina, S., Concetta, R., Francesco, R. & Annalisa, C. SARS-Cov-2 infection: Response of human immune system and possible implications for the rapid test and treatment. Int Immunopharmacol 84, 106519, doi:10.1016/j.intimp.2020.106519 (2020).

18 Zhong, L. et al. Detection of serum IgM and IgG for COVID-19 diagnosis. Sci China Life Sci 63, 777–780, doi:10.1007/s11427-020-1688-9 (2020).

19 Huang, L. et al. Rapid asymptomatic transmission of COVID-19 during the incubation period demonstrating strong infectivity in a cluster of youngsters aged 16–23 years outside Wuhan and characteristics of young patients with COVID-19: A prospective contact-tracing study. J Infect, doi:10.1016/j.jinf.2020.03.006 (2020).

20 Mizumoto, K., Kagaya, K., Zarebski, A. & Chowell, G. Estimating the asymptomatic proportion of coronavirus disease 2019 (COVID-19) cases on board the Diamond Princess cruise ship, Yokohama, Japan, 2020. Euro Surveill 25, doi: 10.2807/1560-7917.ES.2020.25.10.2000180 (2020).

21 Chen, D. et al. Recurrence of positive SARS-CoV-2 RNA in COVID-19: A case report. Int J Infect Dis 93, 297–299, doi:10.1016/j.ijid.2020.03.003 (2020).

22 Zhou, L., Liu, K. & Liu, H. G. [Cause analysis and treatment strategies of "recurrence" with novel coronavirus pneumonia (covid-19) patients after discharge from hospital]. Zhonghua Jie He He Hu Xi Za Zhi 43, E028, doi:10.3760/cma.j.cn112147-20200229-00219 (2020).

23 Sohrabi, C. et al. World Health Organization declares global emergency: A review of the 2019 novel coronavirus (COVID-19). Int J Surg 76, 71–76, doi:10.1016/j.ijsu.2020.02.034 (2020).

